# From CHESS to CHECKMATE: A Practical Score for Predicting Shunt Dependency Following Subarachnoid Hemorrhage

**DOI:** 10.64898/2026.07.18.26358389

**Authors:** Saif Salman, Flora Haidenberger, Mahlika Ahmad, Behnam Rezai Jahromi, Nadia K. Albaramony, Vishal N. Patel, Jeffrey B. Peel, Mutaz Ombada, Salvador F. Gutierrez-Aguirre, Otavio F. de Toledo, Pedro Aguilar-Salinas, Rabih G. Tawk, Richard W. Byrne, Ricardo A. Hanel, Alejandro A. Rabinstein, W. David Freeman

## Abstract

**Objective:** Shunt-dependent hydrocephalus is a common and costly complication of aneurysmal subarachnoid hemorrhage (aSAH), affecting up to 28% of survivors. Existing prediction tools, including the Chronic Hydrocephalus Ensuing from SAH Score (CHESS), have limited discriminative accuracy. We developed the CHECKMATE score, a clinically practical tool to improve prediction of ventriculoperitoneal shunt dependency after aSAH.

**Methods:** In this multicenter retrospective cohort of 486 patients with aSAH from Mayo Clinic (January 1, 2006-December 31, 2021), we used multivariable logistic regression and machine learning to identify independent predictors of ventriculoperitoneal shunt placement. The CHECKMATE score was derived from 5 weighted variables: symptomatic hydrocephalus (10 points), intraventricular hemorrhage (5 points), SAH volume greater than 10 mL (3 points), neutrophil-to-lymphocyte ratio greater than 12 (2 points), and 10-year incremental age thresholds starting at older than 60 years (1 point each).

**Results:** Of 486 patients (mean age, 56.3 years; 64.6% female), 137 (28.2%) required ventriculoperitoneal shunt placement. The CHECKMATE score achieved an area under the curve of 0.808 (compared to 0.737 for CHESS), with a sensitivity of 0.85, specificity of 0.67, and negative predictive value of 0.92 at the optimal cutoff of 14 points.

**Conclusions:** The CHECKMATE score outperforms CHESS for predicting ventriculoperitoneal shunt dependency after aSAH and is easily used at the bedside. Its high negative predictive value helps identify low-risk patients who may benefit from earlier external ventricular drain weaning and shorter hospital stays.

## Introduction

Aneurysmal subarachnoid hemorrhage (aSAH) is a subtype of hemorrhagic stroke with a mortality rate of nearly 40%. Approximately 25% of patients with aSAH die before hospital arrival. Those who survive may experience sequelae that peak around days 4 to 14. Persistent inflammation and scarring of the arachnoid villi make chronic hydrocephalus inevitable.^1^

Cerebrospinal fluid (CSF) shunt placement is a surgical procedure that alleviates persistently elevated intracranial pressure. Patients receiving these shunts, which can be ventriculoperitoneal, ventriculopleural, or ventriculoatrial, risk developing shunt dependency. Approximately up to 30% of patients with aSAH develop shunt-dependent hydrocephalus.^2,3^

CSF shunts impose a financial burden on health care, patients, and their families. The cost of CSF shunt placement per patient is $35,000, in addition to expenses for medical appointments, imaging, and workup. This cost increases when revision surgeries are required for shunt malfunction, displacement, or infections. The 5-year cost can exceed $55,384.66.^4–8^

Several scores that predict shunt-dependent hydrocephalus have been described. Among them, the Chronic Hydrocephalus Ensuing from SAH score (CHESS) has been pivotal for standardized prediction, in addition to other scores that use machine learning (ML)–based approaches.^1,3,9–11^ Herein, we describe our CHECKMATE score, a simple 5-variable tool easily used at the clinical bedside.

## Methods

### Patient Population

This retrospective study was conducted under a protocol approved by the Mayo Clinic Institutional Review Board using deidentified, minimal-risk data. Patients with aSAH admitted to Mayo Clinic in Rochester, Minnesota, and Jacksonville, Florida, between January 1, 2006, and December 31, 2021, were included. Patients without noncontrast computed tomography results, those with traumatic SAH or unconfirmed aneurysm rupture, and those who died in or on arrival at the hospital were excluded.

### Data Management

Demographic, clinical, laboratory, and imaging data were extracted from institutional electronic health records. SAH volume (SAHV) and intracerebral hemorrhage volume were measured on noncontrast computed tomography using the ABC/2 method reported by Föttinger et al.^12^ Symptomatic hydrocephalus was defined as clinical deterioration with ventricular enlargement, and shunt dependency was defined as the need for permanent ventriculoperitoneal shunt placement before discharge. All data management procedures were performed in accordance with institutional privacy and data security standards. Deidentified datasets were used for statistical analysis.

### Statistical Analysis and Score Design

Statistical analyses were performed using Python, version 3.14 (Python Software Foundation). The following libraries were used for preprocessing and classical regression analyses: pandas, NumPy, Matplotlib, statsmodels, and scikit-learn. XGBoost (eXtreme Gradient Boosting), LightGBM (Light Gradient-Boosting Machine; Microsoft Corporation), and CatBoost (Yandex) were used for ML modeling.

Continuous variables were reported as means and standard deviations or medians and interquartile ranges, depending on distribution. Categorical variables were summarized as frequencies and percentages. Numerical features were standardized using the StandardScaler function (scikit-learn developers), and categorical variables were one-hot encoded. Missing numerical values were imputed with the median, while categorical values were imputed with the mode.

A binary logistic regression model was used to identify independent predictors of ventriculoperitoneal shunt dependency after aSAH. The dependent variable was ventriculoperitoneal shunt placement (yes/no). Independent variables included the following demographic, clinical, laboratory, and imaging parameters: age at aSAH, cisternal SAHV, neutrophil-to-lymphocyte ratio (NLR), modified Fisher grade, World Federation of Neurosurgical Societies grade, Glasgow Coma Scale (GCS) score, CHESS, sex, presence of intraventricular hemorrhage (IVH), ventricular enlargement, smoking history, hypertension, diabetes mellitus, alcohol abuse, coronary artery disease, body mass index greater than 30, symptomatic hydrocephalus, intracerebral hemorrhage, and aneurysm location (anterior vs posterior circulation).

Regression coefficients were estimated using the maximum likelihood method and expressed as odds ratios (ORs) with 95% CIs. Variables with *P* values less than .10 in univariable analysis or strong clinical plausibility were retained for multivariable modeling and integration into the ML framework.

To derive a clinically interpretable risk score, significant predictors from the logistic regression were combined with variable importance metrics from advanced ML models (random forest, XGBoost, LightGBM, CatBoost, and support vector machine). This integrative approach provided both statistical interpretability and predictive optimization, balancing model transparency with diagnostic accuracy.

The model’s discrimination was assessed using the area under the receiver operating characteristic curve (AUC-ROC), and performance was further evaluated using accuracy, precision, recall, F1-score, and confusion matrices. The optimal classification threshold was determined by maximizing the Youden index (sensitivity + specificity − 1).

Cutoff values for continuous predictors were defined based on published evidence and exploratory ROC analysis within the present cohort. A SAHV threshold of greater than 10 mL^12^ was adopted from prior volumetric studies linking higher cisternal blood burden to worse outcome. An NLR greater than 12 has been reported as predictive of poor outcomes after aSAH.^13^ For age, a 10-year incremental age threshold starting at older than 60 years was used in line with previous reports showing increased risk of hydrocephalus in elderly patients with aSAH.^14–17^ All tests were 2-tailed, and a *P* value less than .05 was considered statistically significant.

## Results

### Population Characteristics

A total of 486 patients with aSAH were identified. Mean (SD) age at time of aSAH was 56.3 (13.1) years (range, 16-92), and most patients were female (Table 1).

**Table 1.** Baseline Characteristics and Ventriculoperitoneal Shunt Dependency After Aneurysmal Subarachnoid Hemorrhage.

| Characteristic | Total (N=486) |
| --- | --- |
| Sex, No. (%) |  |
| Female | 314 (64.6) |
| Male | 172 (35.4) |
| Age, mean±SD, y | 56.3±13.1 |
| Symptomatic acute hydrocephalus, No. (%) | 312 (64.2) |
| Any CSF diversion (EVD, LD, or LP), No. (%) | 311 (64.0) |
| EVD placed | 237 (48.8) |
| LD placed | 73 (15.0) |
| LP performed | 32 (6.6) |
| Ventriculoperitoneal shunt placement, No. (%) |  |
| Yes (shunt-dependent PHH) | 137 (28.2) |
| No | 346 (71.2) |

Abbreviations: CSF, cerebrospinal fluid; EVD, external ventricular drain; LD, lumbar drainage; LP, lumbar puncture; PHH, posthemorrhagic hydrocephalus.

### Acute Hydrocephalus and Shunt Dependency After aSAH

Symptomatic acute hydrocephalus was found in 312 patients (64.2%). External CSF diversion was performed in 311 patients (64.0%); most underwent external ventricular drainage, followed by lumbar drainage, and lumbar puncture. Ultimately, 137 patients (28.2%) developed shunt-dependent posthemorrhagic hydrocephalus requiring ventriculoperitoneal shunt placement, while 346 (71.2%) did not require a shunt (Table 1).

### Risk Factors for Shunt Dependency and Logistic Regression Model

A multivariable logistic regression analysis was performed to identify independent predictors of ventriculoperitoneal shunt dependency. The model included age at admission, GCS, presence of IVH, alcohol misuse, and symptomatic hydrocephalus.

The model demonstrated a good overall fit (log-likelihood, −215.26; likelihood ratio test, *P*<.001; pseudo-R^2^, 0.25). Age, IVH, and symptomatic hydrocephalus were independent predictors of ventriculoperitoneal shunt dependency, while alcohol misuse showed a nonsignificant trend and GCS was not associated with the outcome.

This regression yielded the best discriminative performance among all tested models, with an in-sample AUC of 0.815 and an internally validated AUC of 0.804±0.027 based on 5-fold stratified cross-validation. This model served as the reference for subsequent analyses integrating additional parameters and for development of the CHECKMATE score (Table 2).

**Table 2.** Multivariable Logistic Regression Model for Predictors of Ventriculoperitoneal Shunt Dependency (Base Model)

| Variable | $\beta$ | SE | z | P value | 95% CI | OR |
| --- | --- | --- | --- | --- | --- | --- |
| Constant | -5.566 | 0.867 | -6.42 | <.001 | -7.27, -3.87 | -- |
| Age | 0.020 | 0.010 | 2.13 | .03 | 0.002, 0.039 | 1.02 |
| GCS | -0.002 | 0.022 | -0.10 | .92 | -0.045, 0.040 | 0.99 |
| IVH | 0.930 | 0.251 | 3.70 | <.001 | 0.44, 1.42 | 2.54 |
| Alcohol abuse | 0.568 | 0.360 | 1.58 | .12 | -0.14, 1.27 | 1.76 |
| Hydrocephalus | 3.465 | 0.601 | 5.76 | <.001 | 2.29, 4.64 | 32.1 |

Abbreviations: GCS, Glasgow Coma Scale; IVH, intraventricular hemorrhage; OR, odds ratio; SE, standard error.

### Expanded Logistic Regression Model Including SAHV and NLR

A second multivariable logistic regression model was developed using the same variables as the initial model, with the addition of SAHV and NLR. The model demonstrated a good overall fit (log-likelihood, −214.72; likelihood ratio test, *P*<.001; pseudo-R^2^, 0.25). Age, IVH, and symptomatic hydrocephalus remained independent predictors of ventriculoperitoneal shunt dependency, while SAHV, NLR, and GCS were not statistically significant. The inclusion of SAHV and NLR slightly reduced overall model discrimination compared to the base model (cross-validated AUC, 0.781±0.023) (Table 3).

**Table 3.**
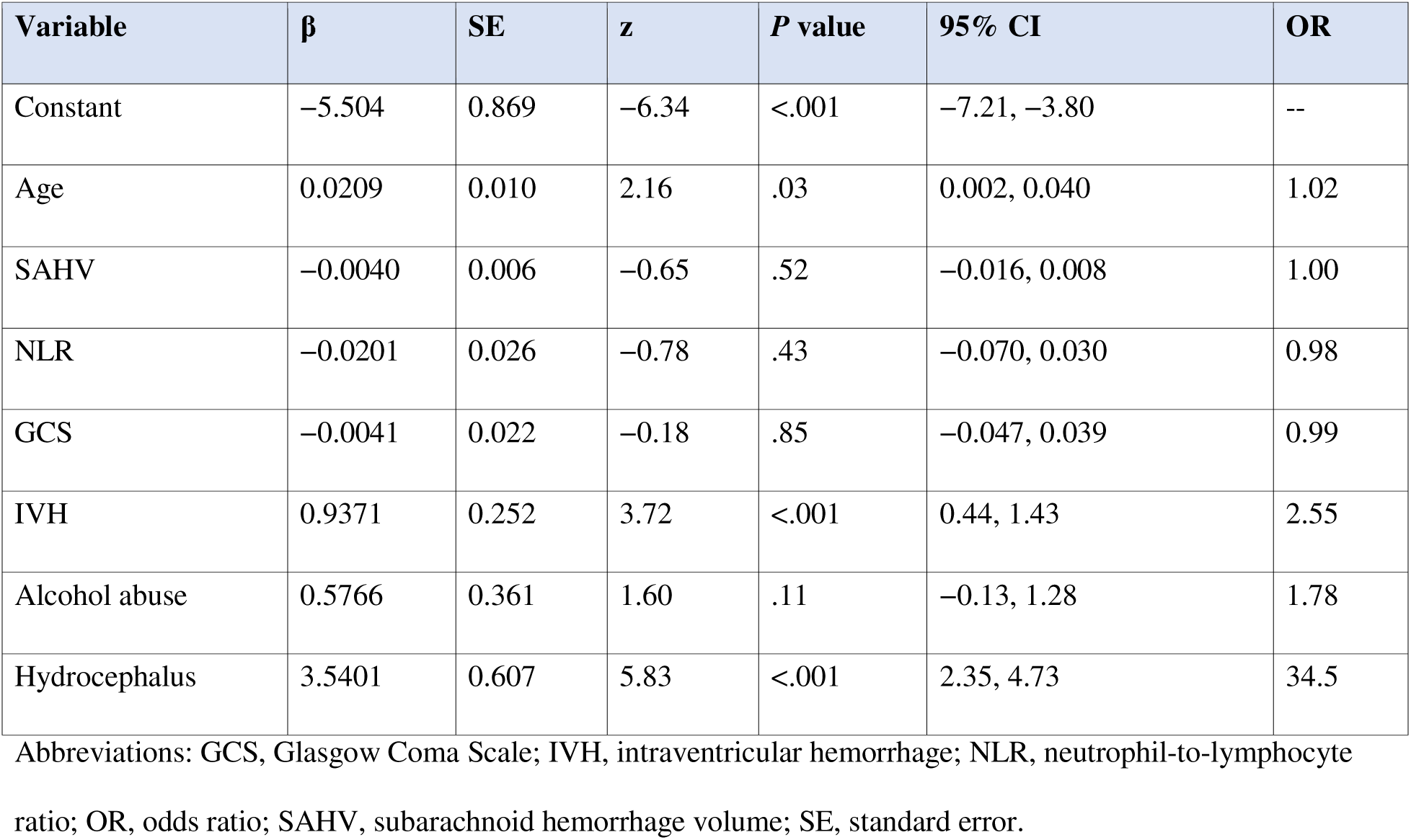
Expanded Logistic Regression Model Including SAHV and NLR.

### Predictive Value of the CHECKMATE Score for Shunt Dependency

The CHECKMATE score was derived from variables identified in the expanded logistic regression model, including age, SAHV, NLR, GCS, IVH, and symptomatic hydrocephalus.

Weighted point values were based on regression effect sizes and adjusted using clinically meaningful thresholds to enhance interpretability and bedside use. Cutoff values for continuous predictors were established from published evidence and exploratory ROC analysis: SAHV greater than 10 mL,^12^ NLR greater than 12,^13^ and age older than 60 years.^14–17^ The final scoring system assigned the following weighted points: symptomatic hydrocephalus, 10 points; IVH, 5 points; SAHV greater than 10 mL, 3 points; NLR greater than 12, 2 points; and 10-year incremental age thresholds (>60, >70, >80, >90 years), 1 point each. In the final cohort of 486 patients, the CHECKMATE score achieved an AUC of 0.808, indicating excellent discrimination for ventriculoperitoneal shunt dependency. The optimal cutoff, determined by the Youden index, was 14 points, corresponding to a sensitivity of 0.85 and specificity of 0.67.

At this threshold, the model yielded a negative predictive value of 0.92, a positive predictive value of 0.50, and an overall accuracy of 0.72. OR analysis demonstrated strong associations between shunt dependency and symptomatic hydrocephalus (OR, 42.9; 95% CI, 13.4-137.3), IVH (OR, 4.93; 95% CI, 3.16-7.69), and ventricular enlargement (OR, 3.93; 95% CI, 2.49-6.21). Additional predictors, including SAHV greater than 10 mL (OR, 2.68; 95% CI, 1.64-4.39) and age older than 60 years (OR, 1.79; 95% CI, 1.20-2.68), remained significant, while NLR greater than 12 showed no significant association (OR, 1.07; 95% CI, 0.46-2.51).

Overall, the CHECKMATE score provides a simple, robust, and clinically applicable tool for predicting shunt dependency after aSAH, maintaining the predictive accuracy of the logistic regression model while outperforming CHESS.

## Discussion

Shunt-dependent hydrocephalus is a long-term complication of aSAH. Despite advances in aneurysm securing, neurocritical care monitoring, and management of delayed cerebral ischemia, permanent CSF diversion remains necessary for some patients. This creates a chronic, resource-intensive condition characterized by device dependence, frequent hospital visits, and substantial health care costs due to shunt revision, infections, and long-term follow-up. This burden highlights the value of a reliable tool to predict the need for permanent CSF diversion upon admission, stratify the risks, and facilitate patient counseling early in the disease course. In this multicenter cohort, we developed the CHECKMATE score, a pragmatic tool for predicting chronic CSF shunt dependency after aSAH. Our aim was to align predictability with clinical decisions while maximizing statistical performance.

CHESS has been pivotal for prediction of shunt dependency after aSAH.^3^ It integrates age, IVH, clinical severity, early infarction, and acute hydrocephalus. The CHECKMATE score builds on CHESS, incorporating 5 variables (age, SAHV, NLR, IVH, and symptomatic hydrocephalus) that are easily collected and standardized across sites. It adjusts predictor weighting and emphasizes variables that directly reflect dysfunction in CSF circulation, such as symptomatic hydrocephalus and IVH. In our cohort, CHESS performance was comparable to previous reports, achieving an AUC of 0.737 with an optimal cutoff of 5 points. However, the CHECKMATE score demonstrated discriminative performance with an AUC of 0.808, while maintaining simplicity and bedside usability (Table 4).

**Table 4.**
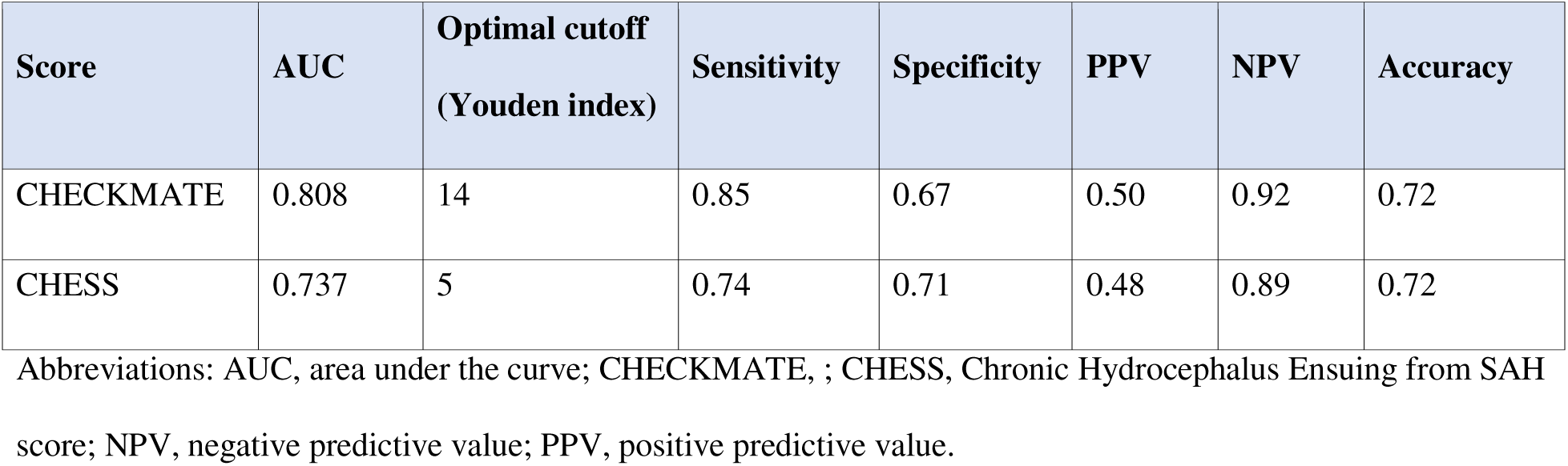
Performance Comparison of CHECKMATE and CHESS Scores for Prediction of Ventriculoperitoneal Shunt Dependency.

The CHECKMATE score has a high negative predictive value (0.92) which supports identification of patients at low risk for chronic shunt dependency. These patients can tolerate earlier weaning of external ventricular drains, resulting in reduced duration of CSF diversion, fewer catheter-related complications, and shorter length of hospital stay.^14–17^

Several ML models demonstrate discriminative performance that exceeds traditional regression-based models in internal validation.^9–11^ They show that shunt dependency is nonlinear and influenced by clinical severity, hemorrhage characteristics, inflammatory response, and treatment approach. Despite having a slightly higher AUC (0.80-0.85), these models rely on complex computations, frameworks, and variables that may not be routinely collected, limiting bedside applicability. For example, Muscas et al^11^ incorporated a set of serial laboratory patterns, radiologic features, and clinical parameters during acute hospitalization. These parameters are time-based and influenced by institution-specific protocols that may vary across sites.^11^ Frey et al^9^ included longitudinal laboratory trajectories, detailed imaging biomarkers, and external ventricular drain management characteristics, such as duration of drainage, drainage volume, and weaning patterns, which may not be consistently documented or standardized across centers.

Gollwitzer et al^10^ incorporated clinical status, change in level of consciousness, need for CSF diversion, and treatment escalation over time, while integrating data from multiple imaging biomarkers from multiple timepoints rather than a single admission scan. Hence, ML models enhance internal accuracy at the expense of real-world feasibility and generalizability when applied externally.^18,19^

Another methodological consideration highlighted by our findings is the distinction between biologic relevance and independent predictive values. ML models can assign high importance to certain variables, such as serial inflammatory markers, during training. However, once downstream clinical states such as IVH and symptomatic hydrocephalus are accounted for, the predictive contribution of some upstream values diminishes. Therefore, without careful clinical oversight, there is a risk of overemphasizing factors that are limited by techniques, timing, sequence, and institutional practices.

We designed the CHECKMATE score to address these limitations by prioritizing downstream manifestations of impaired CSF dynamics that directly translate to bedside decision-making. Although we initially used ML models to explore development and interaction features, the final model evolved into a transparent additive score grounded in clinical reasoning and pathophysiologic coherence. Hence, the CHECKMATE score serves as a predictive tool at patient admission and can be integrated into more advanced multimodal artificial intelligence systems.

We included factors such as GCS, NLR, and SAHV in the expanded CHECKMATE score. The rationale for including SAHV was based on its association with delayed cerebral ischemia and mortality.^12,20,21^ The rationale for including NLR was based on the role of inflammation in secondary injury after aSAH.^13,22,23^ However, the expanded logistic regression model showed slightly reduced overall discrimination compared to the base model (cross-validated AUC, 0.781±0.023 vs 0.804). This can be attributed to the SAHV values in our cohort, which ranged from 0.0 to 253.9 mL, indicating wide interindividual variability in subarachnoid blood volume. Furthermore, some patients had missing NLR values as differential blood counts were not consistently available on the day of admission.

Despite these limitations, both SAHV and NLR were retained because they are objective, quantitative parameters with potential for automated extraction and real-time computation in future imaging-based prediction workflows. Further studies are needed for validation, as we still believe in the mechanistic link between NLR, SAHV, and shunt dependency.

This study has several limitations. As a retrospective analysis, it reflects real-world clinical practice and may be subject to inherent biases. Although derived from a multicenter cohort, variability in institutional practices should be considered. Furthermore, the model was internally validated, and external validation in independent cohorts is needed to confirm its broader applicability.

## Conclusion

The CHECKMATE score is easily incorporated into current practice, while the expanded CHECKMATE serves as an exploratory bridge toward future multimodal artificial intelligence systems.

**Figure 1.**
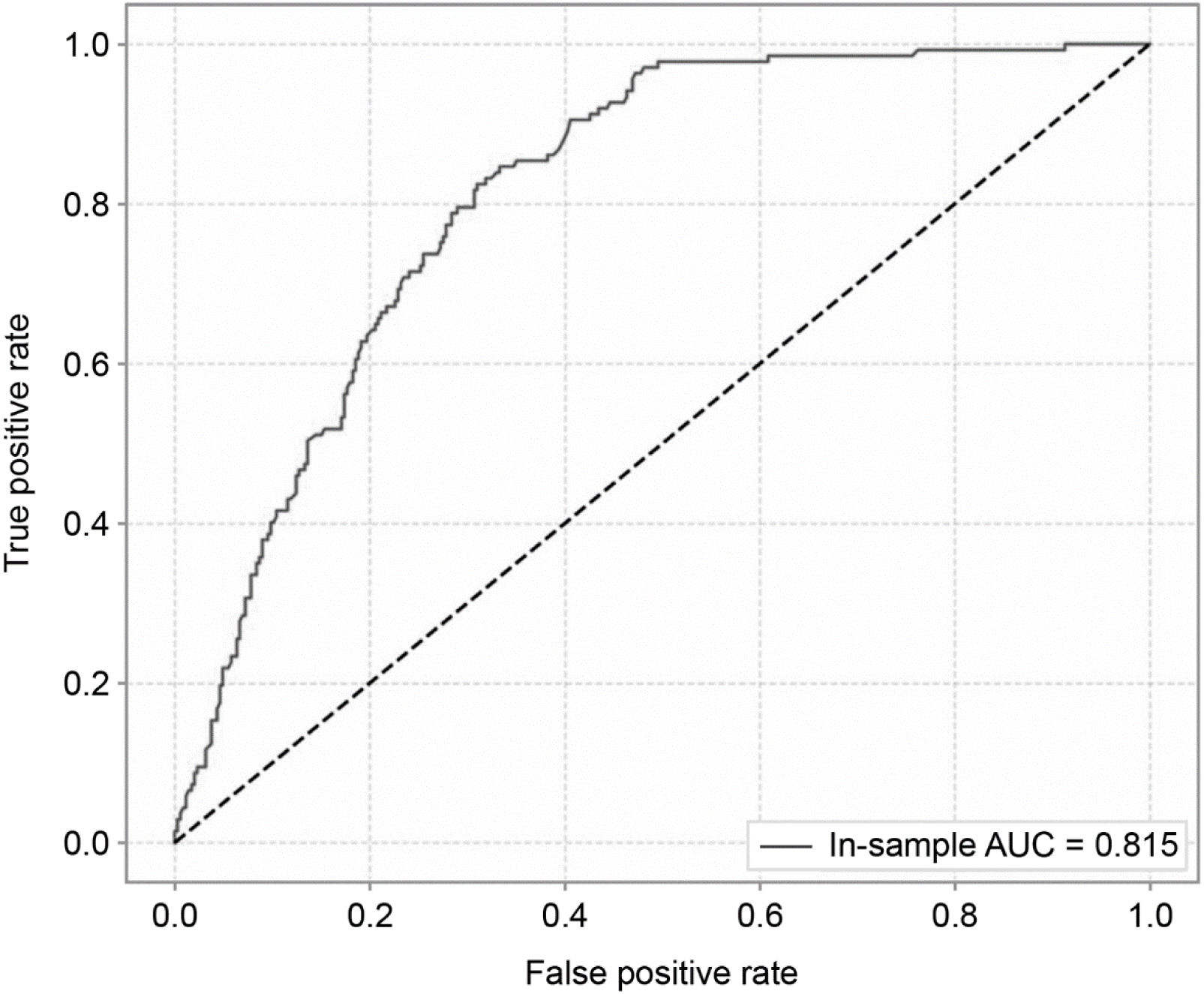
ROC Curve of the Multivariable Regression Model of Shunt-Dependency Predictors. AUC indicates area under the curve; ROC, receiver operating characteristic curve.

**Figure 2.**
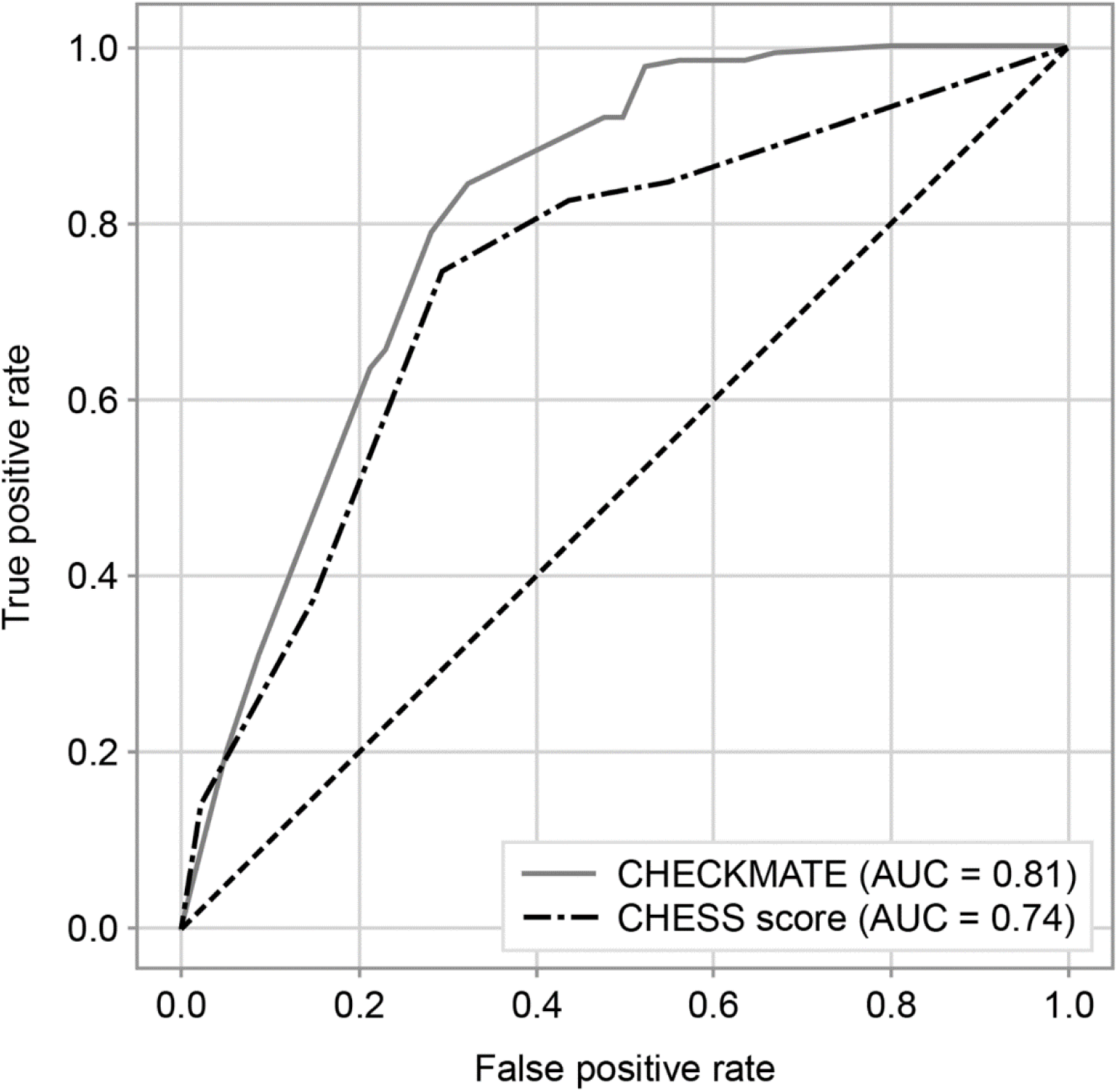
ROC Curve of CHECKMATE vs CHESS Score. AUC indicates area under the curve; CHESS, Chronic Hydrocephalus Ensuing from SAH score; ROC, receiver operating characteristic curve.

## Data Availability

All data produced in the present study are available upon reasonable request to the authors

## Acknowledgment

We thank our AI Engineers in the SAHVAI Lab: Jordan Walker, Mark Roberts, and Ryan A. Rowe, and the Google Advanced Solutions Lab: Sanjanna Reddy, PhD, and Benoit Dherin, PhD. The Scientific Publications staff at Mayo Clinic provided copyediting, proofreading, administrative, and clerical support.

